# Performance Characteristics of Reasoning Large Language Models for Evidence Extraction from Clinical Genomics Literature

**DOI:** 10.64898/2026.02.18.26346543

**Authors:** Mullai Murugan, Bo Yuan, Joshi Stephen, Charul Gijavanekar, Shengfeng Xu, Senkottuvelan Kadirvel, E. Andres Rivera-Munoz, Vitor Manita, Francisco Delca, Richard A Gibbs, Eric Venner

**Author notes:** Corresponding Author: Mullai Murugan.

## Abstract

**BACKGROUND:** Genetic variant curation requires literature-guided comparison of variant prevalence in affected individuals versus healthy controls. This evidence is categorized as the PS4 evidence code by the AMP/ACMG variant interpretation guidelines and its manual extraction is a major bottleneck in clinical variant curation. We evaluated whether reasoning-capable large language models (LLMs) can support guideline-constrained PS4 evidence extraction from literature.

**METHODS:** We benchmarked twelve reasoning-capable LLMs spanning providers, capability tiers, and releases on 281 publication–variant pairs against an expert-curated truth-set. Models performed variant detection (Task 1) and PS4-eligible proband counting under ACMG/AMP and gene-specific ClinGen Variant Curation Expert Panel guidance (Task 2). Six newer models also underwent variant-level PS4 strength assignment (Task 3). We compared guideline-informed and minimal baseline prompts, categorized errors by blinded dual review, and assessed model performance, reproducibility, cost, and latency.

**RESULTS:** Task 1 concordance was 93.2-99.6%, with 10 of 12 models exceeding 96%. Task 2 exact-count concordance was 72.2-92.2%, and Task 3 was 71.4-92.9%. Removing guidance minimally affected Task 1 but reduced Task 2 concordance by 14.6-33.1 points in all six newer models, with the largest losses in lower-performing baseline models. Variant-mapping errors decreased in newer releases, whereas phenotype-eligibility and case-independence errors persisted. Five of six newer models were statistically indistinguishable on Task 2; with only the smallest model lagging. Among those five, cost varied 6.9-fold and latency 2.9-fold.

**CONCLUSIONS:** Reasoning-capable LLMs can support automation of guideline-based PS4 evidence extraction, achieving high concordance with expert curation. Performance depends on supplying relevant guidelines and errors more often involve guideline application than evidence identification. Residual phenotype and case-independence errors support a hybrid workflow with expert review, while statistically comparable performance among five newer models suggests that cost, latency, and availability may be relevant considerations when selecting among similarly performing models.

## Introduction

Interpreting the clinical significance of genetic variants is central to genomic diagnosis and research. The ACMG/AMP guidelines^1^ provide a standardized framework for variant classification through evidence codes derived from multiple sources, including population data, functional studies and clinical observations. Among these, the PS4 criterion, which supports pathogenicity when a variant is enriched in affected individuals compared with controls, is amongst the most useful of classifiers. Applying PS4 is extremely labor-intensive, however, requiring manual synthesis of evidence from case reports and cohort studies, and often adjudications of phenotype relevance and family structure. As population-scale sequencing initiatives (e.g., *All of Us*)^2^ expand, this manual process increases interpretive variability and limits scalability.

Although Large language models (LLMs) have been explored across clinical and biomedical applications^3–14^ including variant interpretation^15–19^, PS4 poses a distinct challenge, the case and variant information needed is dispersed across publication texts, tables, figures and supplementary materials. Scaling this requires new approaches beyond manual review to reconcile case details and determine eligibility. Recent advances in reasoning-capable LLMs are designed to support complex, multistep problem solving through approaches such as reinforcement learning-based optimization, advanced prompting strategies, and self-correction^20–23^, leading to the hypothesis that these models might be able to address the PS4 problem.

To test this, we developed an LLM-based framework to automate the extraction of PS4 evidence from biomedical publications under ACMG/AMP and ClinGen Variant Curation Expert Panel (VCEP) guidance^24^ and we benchmarked contemporary reasoning-capable LLMs using an expert-curated truth-set. We evaluated models on two tasks: (1) determining whether a prespecified variant was reported in the publication and (2) when present, counting independent PS4-eligible probands under the applicable guidance. In an exploratory third task (3), we asked whether a model could aggregate its own per-publication output across every publication reporting a variant and assign that variant’s overall PS4 strength based on VCEP guidance. We evaluated truth-set concordance, error modes, prompt sensitivity, cost, latency and reproducibility to characterize the current utility and limitations of reasoning-capable LLMs for guideline-driven evidence extraction. By reducing manual workload and improving reproducibility, this approach could help scale variant interpretation pipelines to meet the growing demand for clinical genome analysis.

## Methods

### Truth-Set Construction

We assembled an expert-curated truth-set of biomedical publications to evaluate automated PS4 evidence extraction. Publications (including supplements) were drawn from historical variant-interpretation records at the HGSC Clinical Laboratory (HGSC-CL) and the ClinGen Evidence Repository (https://erepo.clinicalgenome.org/evrepo/). The truth-set comprised 281 publication-variant pairs drawn from 275 peer-reviewed publications, spanning 58 genes and 128 distinct variants. The dataset included both true-positive and true-negative cases (Supplementary Tables S1, S2). Negative controls were created by starting from and actual publication-variant pair and applying one of a set of standard transformations: replacing all variant information with that of a variant from a different gene, changing the variant nucleotide in the cHGVS, changing the reference nucleotide in the cHGVS, entirely replacing the cHGVS but not the pHGVS, modifying both the pHGVS, and cHGVS, or modifying the transcript without changing the pHGVS or cHGVS.

For each publication-variant pair, curators recorded two ground-truth labels: (1) whether the publication reported the target variant (Task 1) and (2) the number of independent PS4-eligible probands with phenotypes consistent with the gene-disease context attributable to that variant, applying ACMG/AMP PS4 criteria and ClinGen VCEP guidance when applicable (Task 2). As a limited proof of concept (PoC), for a secondary subset of 91 publication–variant pairs representing 14 variants in 13 genes, curators also integrated evidence across publications and assigned variant-level PS4 strength (Task 3). Records also captured bibliographic identifiers, standardized variant descriptors (gene, transcript, HGVS cDNA and protein nomenclature, and genomic coordinates), and curated clinical context (e.g., reported phenotypes and inheritance).

### Model Selection

We evaluated a panel of twelve reasoning-capable LLMs across two evaluation phases. Phase 1 evaluated six models released in 2025: Google Gemini 2.5 Pro, OpenAI GPT-5, OpenAI o3, OpenAI o4-mini, Anthropic Claude Opus 4.1, and Anthropic Claude Sonnet 4. Phase 2 evaluated six successor and comparison models released through mid-2026: Google Gemini 3.1 Pro Preview, Anthropic Claude Opus 4.8, Anthropic Claude Sonnet 5, Anthropic Claude Haiku 4.5, OpenAI GPT-5.6 Sol and Z.ai GLM-5. Together, the 12-model panel represents multiple providers, capability tiers (frontier, mid-tier, small), reasoning architectures, and open-weight access (GLM-5), including four same-family version pairs. All models were evaluated under identical prompt and context configuration on the 281-pair truth set; model identifiers and configurations are detailed in Supplementary Table S1.

### Pipeline Design

We implemented a three-part, guideline-informed variant interpretation pipeline for PS4 criteria using direct model API invocations (Figure 1):

**Figure 1:**
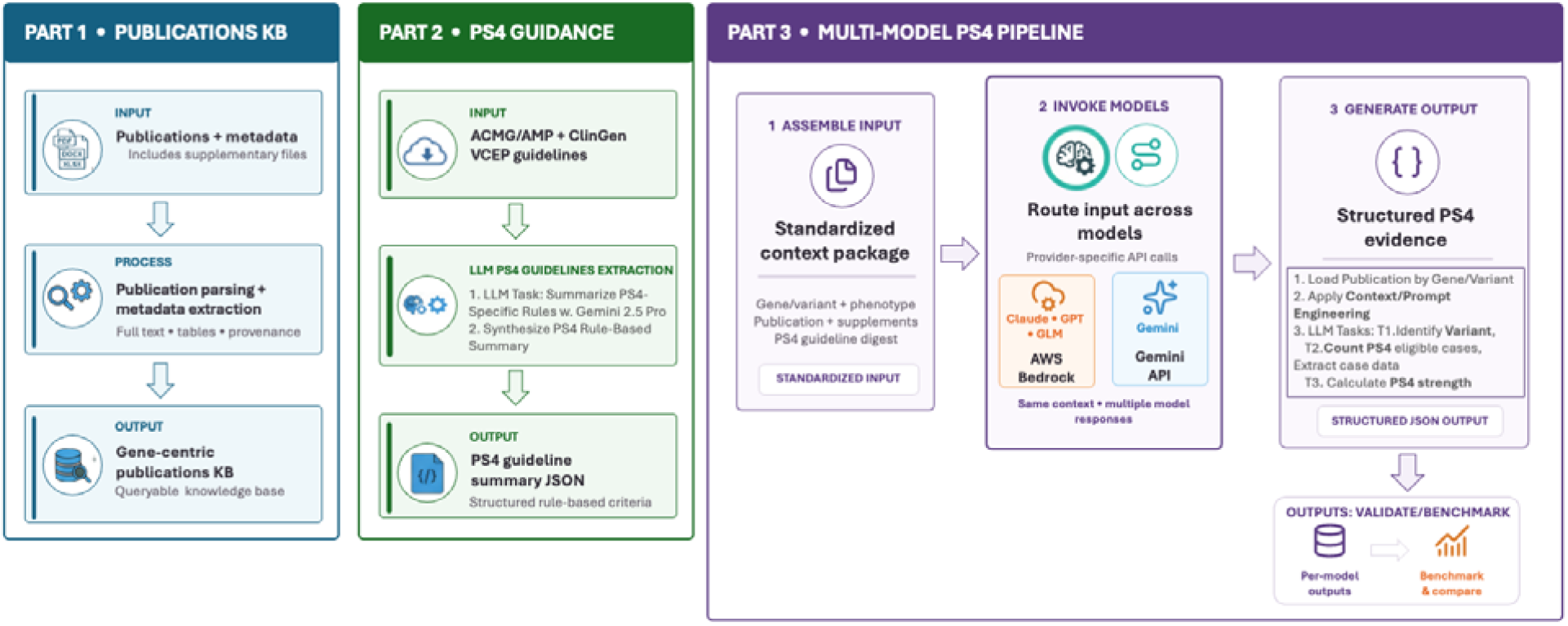
End-to-End Pipeline for PS4 Evidence Extraction and Case Counting. Schematic overview of the three-stage workflow. Part 1 (Publications knowledge base (KB) creation): Publications and associated metadata (including supplements) are parsed and indexed to create a queryable knowledge base. Part 2 (PS4 Guideline summarizer): ACMG/AMP and ClinGen VCEP guidance are summarized into PS4-specific, rule-based criteria and exported as a structured JSON to support consistent, guideline-constrained extraction using Gemini 2.5 Pro LLM. Part 3 (PS4 Pipeline): Orchestrates per publication-variant processing by generating standardized prompts for Task 1 (T1) variant detection, Task 2 (T2) PS4-eligible case counting and Task 3 (T3) PS4 strength assessment, invoking each LLM with these prompts, and producing structured evidence summaries. Model outputs are written to separate, schema-specific CSV files for each task to support downstream evaluation and analysis.

#### Part 1 - Literature knowledge base (KB)

All truth-set publications and supplements were parsed to plain text and organized into a gene-centric KB to standardize model inputs. Each entry retained bibliographic metadata and file provenance to support traceability of extracted evidence to source documents (Figure 1, Part 1).

#### Part 2 - Guideline summarization (PS4 digests)

We generated PS4 digests from ACMG/AMP and gene-specific ClinGen VCEP guidance when available. Using Gemini 2.5 Pro, each digest encoded PS4 mode (proband-count vs case-control), permitted phenotypes, case-independence rules, and inheritance constraints, with each rule mapped to cited source text. Digests were cached per gene, with fallback to generic ACMG PS4 when gene-specific guidance was unavailable (Figure 1, Part 2).

#### Part 3 – Model Invocation and Context Management

All primary benchmark results were generated via direct model API invocations (Figure 1, Part 3). For each publication-variant pair, pipeline code invoked models with a prompt containing the target gene-variant, phenotype context, gene-specific PS4 guideline digest, and publication full text. Rather than using retrieval-augmented generation^25–27^, text summarization^28,29^, or agentic orchestration^30,31^ - which can introduce variance between models - we supplied the entire publication and supplementary text directly within each model’s input context.

Models processed each publication end-to-end to return structured JSON outputs for three tasks: variant detection (Task 1), PS4-eligible proband counting under the gene-specific digest (Task 2), and variant-level PS4 strength aggregated across publications (Task 3).

Outputs included the variant-detection call, PS4 case count, PS4 strength assessment, supporting rationale and source citations. For comparability, input context was capped at 200,000 tokens, the lowest limit among the evaluated models, which accommodated all 281 pairs. Full prompts and output schemas are in Supplementary Text S1.

#### Prompt design

To estimate the contribution of domain-specific guidance beyond the models’ pretrained knowledge, we compared the guideline-informed configuration (v7) with a minimal baseline (v0) across the same 281 pairs and six Phase 2 models (Supplementary Text S1). Both received identical gene, transcript, variant, phenotype and parsed publication context. The baseline asked only whether the publication reported the variant and how many reported cases were PS4-eligible based on the model’s existing knowledge of ACMG/AMP and ClinGen guidance. The v7 configuration additionally supplied ACMG/AMP and gene-specific ClinGen PS4 guidance, phenotype and inheritance eligibility rules, study-design distinctions, and case-independence and relatedness instructions. Model versions, inference settings, context assembly and limits, output schema and scoring were otherwise held constant.

#### Token Usage, Latency, and Cost Accounting

We tracked input tokens, output tokens and wall-clock latency for the Phase 2 models on all 281 pairs and computed per-pair costs from recorded token volumes using published on-demand list rates in force at the time of evaluation. All models ran on AWS Bedrock (*us-east-1*) except Gemini 3.1 Pro Preview, which was unavailable there and ran on the Gemini API at Google Console list pricing.

#### Negative-control (hallucination) analysis

To evaluate the models’ propensity to hallucinate or falsely report the presence of variants in a publication, we generated 28 negative-control pairs in which the queried variant specification (HGVSc, HGVSp, HGVSg, and/or transcript) was either fictional or a real variant known to be absent from the publication (Supplementary Table S2). These pairs were processed using the same pipeline for each model (identical inputs, prompt, and output schema), and Task 1 variant-detection calls were manually compared with curator adjudication.

#### Evaluation metrics and statistical analysis

Tasks 1 and 2 were evaluated against the curated truth set on the same 281 publication–variant pairs for every model. For Task 1 we report accuracy, sensitivity and specificity. For Task 2 we report exact-count concordance, the proportion of pairs whose predicted PS4 case count matched the truth-set count. Task 3 was evaluated against the curated variant-level strength tier, reporting exact tier agreement and the ordinal distance and direction of each disagreement. Proportions carry 95% Wilson confidence intervals. All paired comparisons use exact McNemar tests, Holm-corrected. Two reviewers independently assigned each Task 2 discordance to one of 11 prespecified error categories. We report observed agreement and Cohen’s κ, and disagreements were resolved by a third reviewer blinded to the identities of the initial reviewers. Code and data on GitHub^32^.

## Results

### Model performance across Task 1, Task 2 and Task 3

Model performance and concordance with the truth-set varied across tasks. **Task 1**, detecting a pre-specified variant, was near ceiling: truth-set concordance across the 281 publication-variant pairs ranged from 93.2% to 99.6%, and ten of twelve models exceeded 96% (Figure 2.A, Supplementary Table S3). Variant representation accounted for most failures, with eleven of twelve Claude Sonnet 4 false negatives involving legacy aliases or alternate numbering (Supplementary Example S1). **Task 2**, counting PS4-eligible cases, requires guideline execution and was harder and more variable, ranging from 72.2% to 92.2% (Figure 2.B). The spread across models was 20.0 percentage points for counting compared to 6.4 for detection. **Task 3** concordance with the truth-set, assigning variant-level PS4 strength by aggregating across every publication reporting an eligible case, ranged from 71.4% to 92.9%, highest for Claude Sonnet 5 and for Claude Opus 4.8 with extended thinking enabled (Figure 2.C, 2.D). Two causes predominated: failure to de-duplicate probands across publications, as for LDLR c.1195G>A (Gemini 3.1 Pro) and GCK c.449T>A (GLM-5); and missing thresholds, as for RYR1 c.7291G>A, where every model recovered the correct count of two probands but four assigned Supporting rather than the curated Moderate.

**Figure 2:**
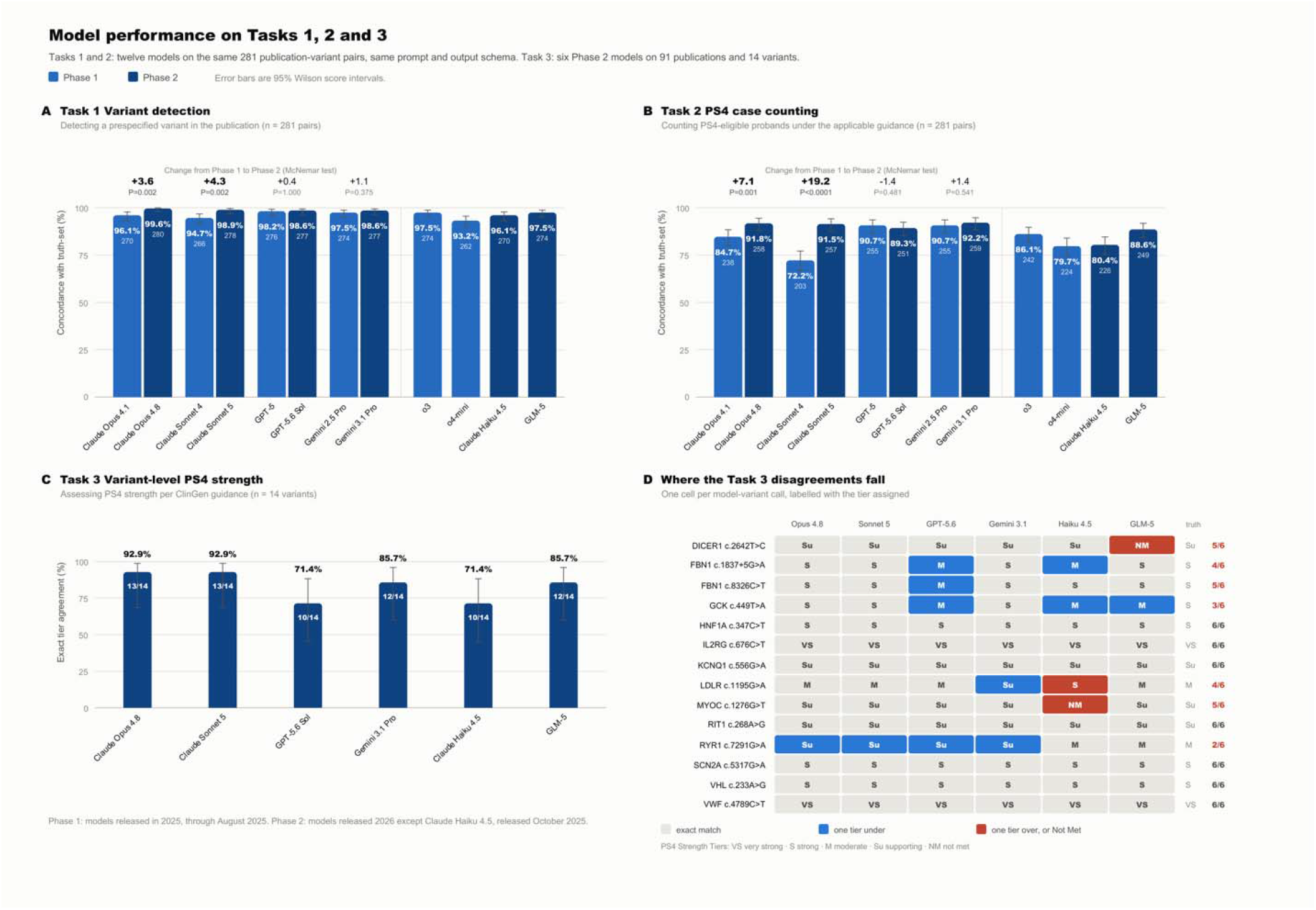
Model performance across Tasks 1, 2 and 3. **(A) Task 1, variant detection** and **(B) Task 2, PS4 case counting.** For each of the twelve models, bars show concordance with the expert-curated truth-set across the same 281 publication-variant pairs, evaluated with the same prompt and output schema. Task 1 concordance is the proportion of pairs whose variant presence call matched the truth-set; Tsk 2 concordance is the proportion whose PS4 case count matched exactly, scored correct only when the model also established variant presence correctly. Each bar is labeled with its percentage and, beneath it, the number of pairs concordant; the denominator is constant within a panel and is given in the panel subtitle. Light bars are Phase 1 models (released through August 2025) and dark bars Phase 2 (from October 2025). The four model families with a release in both phases are drawn as adjacent pairs, with the change in percentage points and the exact McNemar P value shown above each pair; the four models with no counterpart in the other window follow to the right of the divider and carry no paired comparison. On Task 2 the paired changes were 84.7% to 91.8% for Claude Opus 4.1 to 4.8 (P=0.0008), 72.2% to 91.5% for Claude Sonnet 4 to 5 (P<0.001), 90.8% to 89.3% for GPT-5 to GPT-5.6 Sol (P=0.48), and 90.8% to 92.2% for Gemini 2.5 Pro to 3.1 Pro (P=0.54). On Task 1 both Claude lineages also improved significantly (P=0.002 and P=0.0018) while the GPT and Gemini lineages did not. Error bars are 95% Wilson score intervals. **(C) Task 3, variant-level PS4 strength.** For each of the six Phase 2 models, bars show agreement with the curated truth-set strength tier across 14 variants. Percentages are shown above each bar with the number of variants concordant inside. **(D) Per-variant detail for Task 3.** One cell per model-variant call, labeled with the strength tier the model assigned (VS very strong, S strong, M moderate, Su supporting, NM not met). Cell color encodes the direction of disagreement with the curated tier, shown at right, and the final column gives the number of models assigning the correct tier for that variant.

Four families had a release in both phases (Figure 2.A, 2.B). Improvement was confined to the lineages that started lowest: both Claude lineages gained significantly on Task 2, Opus from 84.7% to 91.8% (P=0.0008) and Sonnet from 72.2% to 91.5% (P<0.001), whereas GPT and Gemini, already at 90.8%, did not change. Inference configuration mattered as much or more: raising GLM-5’s reasoning effort improved case counting by 6 percentage points (P=0.021), a larger gain than most release changes we measured (Supplementary Text S4).

### Nature of Task 2 model errors

Two reviewers independently categorized every Task 2 discordance on a closed eleven-category taxonomy (Supplementary Text S3), covering all six Phase 1 models and one lineage per vendor for Phase 2 (Figure 3.A, 3.B). Agreement was 71.9% for Phase 1 (Cohen’s kappa 0.66, n=267) and 46.7% for Phase 2 (kappa 0.40, n=75). A blinded third reviewer adjudicated all 115 disagreements, favoring neither reviewer in Phase 1 (binomial P=0.80) and neither significantly in Phase 2 (P=0.12). Nineteen rows were ruled to require truth-set revision rather than a model-error category, leaving 323 categorized model errors (Figure 3.C).

**Figure 3:**
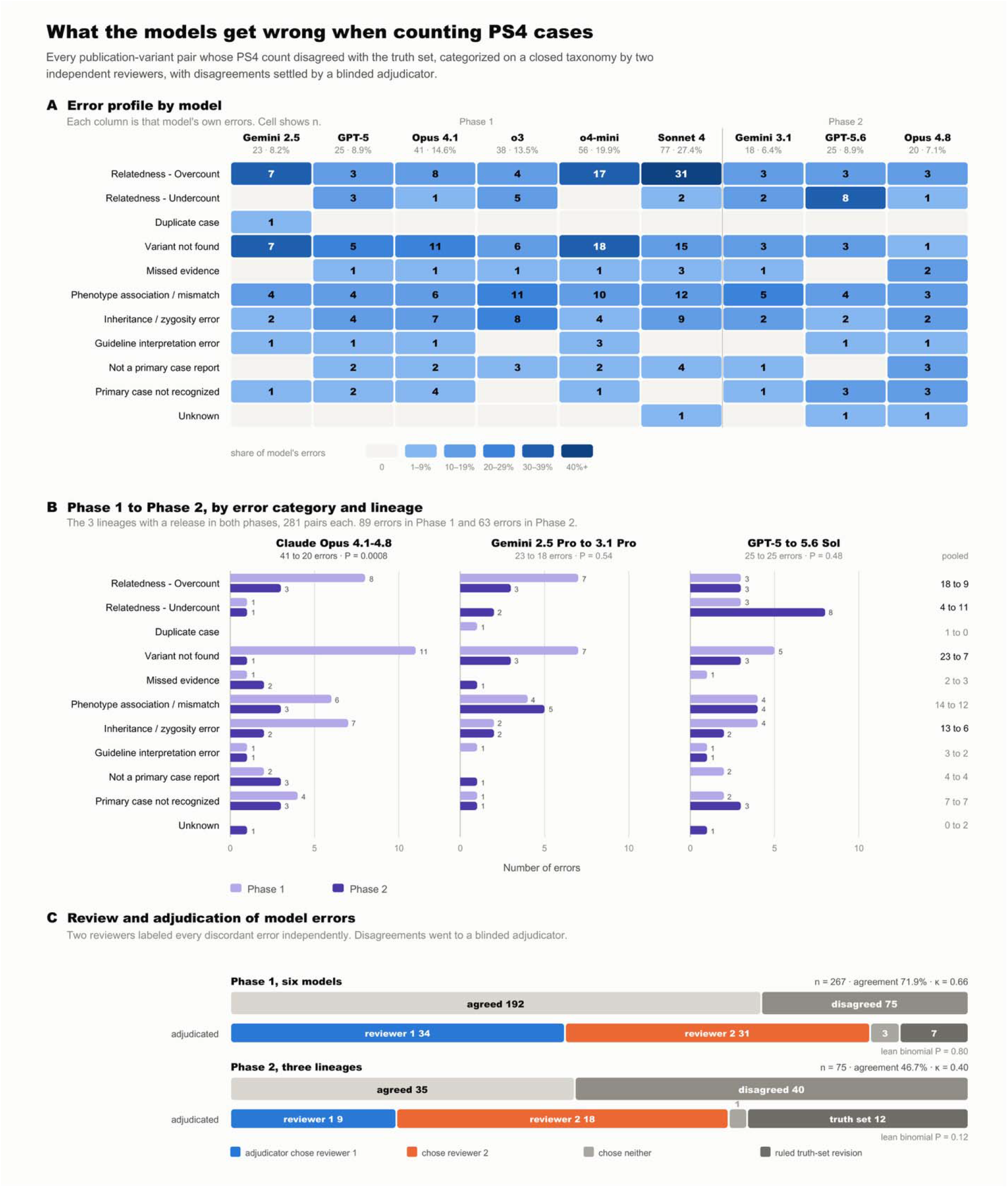
Task 2 (PS4 case count extraction): Model performance and error patterns. Every publication-variant pair whose PS4 case count disagreed with the truth set was assigned one category from a closed eleven-category error taxonomy by two reviewers working independently. Categorization covers all six Phase 1 models and the Phase 2 releases of the Claude Opus, Gemini and GPT lineages. Rows the adjudicator ruled to require truth-set revision rather than a model error are excluded from panels A and B and reported in panel C. **(A) Error profile by model**. Cells give the number of errors in each category, shaded by that category’s share of the model’s own errors, so profiles are comparable across models that differ in overall accuracy. Column headers give each model’s total errors and its error rate over the 281 scored pairs. Categories: **Relatedness - overcount**, related individuals counted as independent probands; **Relatedness - undercount,** independent cases treated as possibly related; *Duplicate case*, the same individual counted twice across reports; **Variant not found**, the prespecified variant not identified or incorrectly mapped; **Missed evidence**, information present in the publication but missed by the model; **Phenotype association / mismatch**, cases included or excluded against the PS4-eligible phenotype; **Inheritance / zygosity error**, zygosity or inheritance context misclassified; **Guideline interpretation error**, the PS4 mode or its thresholds misapplied; **Not a primary case report**, a review or functional study treated as case evidence; **Primary case not recognized**, genuine case evidence not counted; **Unknown**, not attributable to one category. **(B) Error profile changes from Phase 1 to Phase 2**. Error profiles across Phase 1 and Phase 2 lineages for Claude Opus, Gemini and GPT. Lineage headers give total errors and the exact McNemar P value for the overall change; pooled per-category counts are at right. 89 Phase 1 to 63 Phase 2 errors, distributed across eleven error categories. **(C) Review and adjudication of model errors.** Two reviewers independently categorized Task 2 discordance for each error, covering all six Phase 1 models and, for Phase 2, one lineage per vendor (Claude Opus, Gemini and GPT). Agreement was 71.9% for Phase 1 (Cohen’s kappa 0.66, n=267) and 46.7% for Phase 2 (kappa 0.40, n=75). A blinded third reviewer adjudicated all 115 disagreements, favoring neither reviewer in Phase 1 (binomial P=0.80) and neither significantly in Phase 2 (P=0.12). Nineteen rows were ruled to require truth-set revision rather than a model-error category, leaving 323 categorized model errors.

In Phase 1, two mechanisms accounted for half of all errors: non-conservative counting of related individuals (70, 26.9%) and failure to locate or map the variant (62, 23.8%) with relatedness overcounting accounted for 31 of Claude Sonnet 4’s 77 errors (Supplementary Example S3). The two categories, Relatedness-overcounting and Duplicate case were asymmetrical: related individuals were counted as independent probands in 79 of the 323 adjudicated errors, whereas the same individual was counted twice in only one (Supplementary Example S4), indicating that models conflate family structure far more often than they duplicate a single case.

For Phase 2 (Figure 3.B), variant-not-found errors fell from 23 to 7 in all three, whereas phenotype mismatch and the two case-ascertainment categories did not change. Relatedness changed in character rather than in volume: over-counting fell from 18 to 9 while under-counting rose from 4 to 11, reversing the Phase 1 asymmetry, and that rise was almost entirely GPT. Only the Claude Opus lineage reduced its total significantly (41 to 20, P=0.0008).

### Contribution of guideline-informed prompting

We estimated the combined contribution of the instructions and the injected PS4 guidance by comparing the v7 prompt against a minimal unaided baseline (v0) on the same publication-variant pairs, models and inference configuration, for Tasks 1 and 2 on the Phase 2 models (Figure 4). For task 1, primarily semantic extraction, was unaffected: −1.4 to +0.4 percentage points, none significant (P≥0.25; Figure 4.A). Task 2 requires executing ACMG/AMP and ClinGen eligibility and independence rules, and every model lost 14.6 to 33.1 points under v0 (mean 24.7), all significant (Figure 4.B). Guidance benefited the weakest models most and compressed the field: the gain scaled inversely with unaided performance (Spearman rho = −0.94, P=0.005), between-model spread fell from 27.4 to 11.8 points, and ranking was not preserved. The loss was directional, with models over-counting two to four times more often than under-counting.

**Figure 4:**
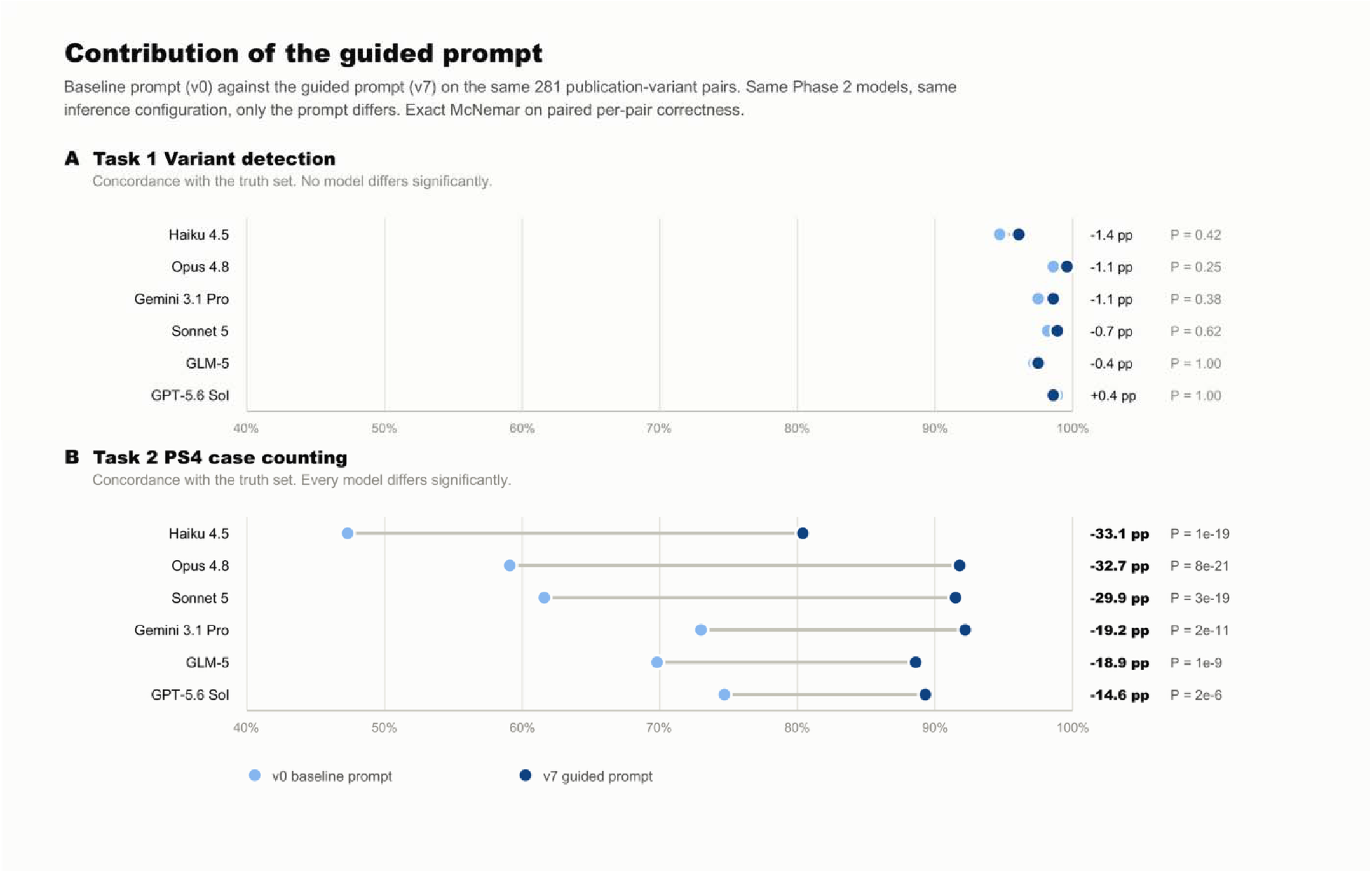
Contribution of guideline-informed prompting. For each of the six Phase 2 models, concordance with the expert-curated truth-set under a minimal baseline prompt (v0, light blue) and the guideline-informed prompt (v7, dark blue), evaluated on the same 281 publication-variant pairs with the same models and inference configuration; only the prompt differs. The v0 prompt supplies the gene, transcript, variant, phenotype and publication text but omits the gene-specific PS4 specification, the eligibility and case-independence rules and counting guidance (see Methods). The line connecting each pair of points spans the change. **(A) Task 1, variant detection. (B) Task 2, PS4 case counting.** Models are ordered by the size of that change, which is given in percentage points to the right of each pair together with the P value from an exact McNemar test on paired per-pair correctness.

Three genes whose VCEP guidelines do not evaluate PS4 by proband counting (MLH1, MSH6, PMS2; 20 publication-variant pairs) illustrate this. Using both prompts, every model found the variant in all 20 test cases. This indicates that the difference is guidance rather than retrieval: v0 returned 278 cases against a truth of zero with no model invoking the specification, v7 correctly returned zero throughout (Supplementary Example S5).

### Token usage, latency and cost

Input and output token usage was recorded for all Phase 2 models (Figure 5.A), ranging from 12.70 million tokens (Claude Sonnet 5) to 7.95 million (GPT-5.6 Sol). Costed at published on-demand rates, the benchmark ranged 6.9-fold, from $12.06 (GLM-5) to $82.95 (Claude Opus 4.8), or $0.043 to $0.295 per publication–variant pair. Mean latency ranged from 29.1 seconds per pair (Gemini 3.1 Pro) to 85.3 seconds (GLM-5). Among the five models that could not be separated on Task 2, Gemini 3.1 Pro was fastest and GLM-5 cheapest at roughly a third the cost of any other in the group (Figures 5.B, 5.C). Concordance with truth-set did not distinguish these five, leaving selection to cost and latency.

**Figure 5:**
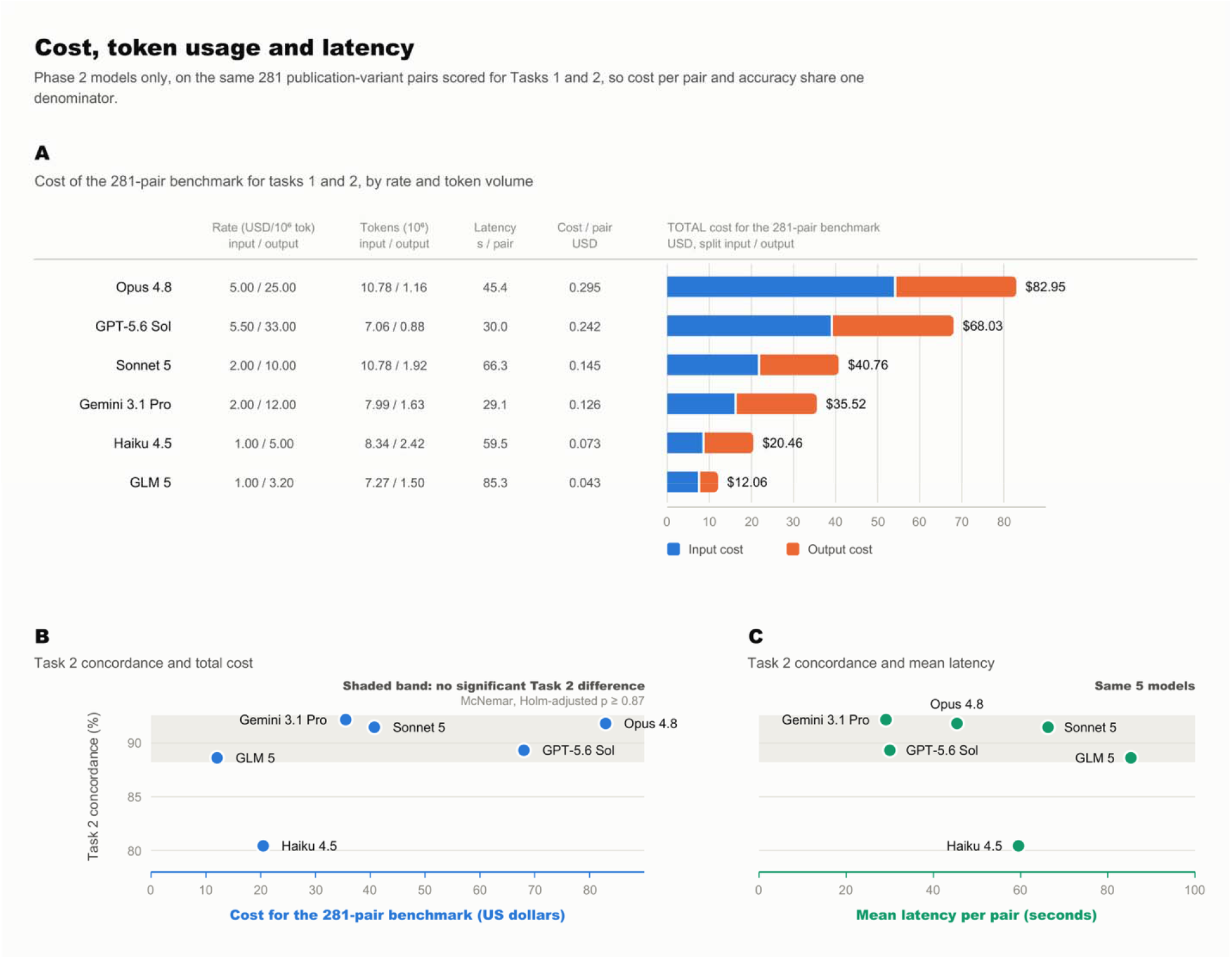
Token usage, latency and cost for the Phase 2 models (N = 281 pairs). (A) Published on-demand rate, token volume, mean latency and cost per pair for each model. Bars show TOTAL benchmark cost split into input and output. Input counts differ across models on identical text because tokenizers differ. (B) Task 2 concordance against total cost and (C) against mean latency. The shaded band spans the five models not statistically separable on Task 2 (exact McNemar, Holm-adjusted P ≥ 0.64), within which cost varies 6.9-fold. Costs use published rates, not billed amounts.

### Analysis of negative controls

Across 28 negative-control pairs (Supplementary Table S2), models correctly reported the queried variant was absent in 22(78.57%), including pairs where only part of the variant description had been modified. Importantly, positive calls were not hallucinations; they reflected partial matching to an unchanged element of the query rather than requiring concordance across the HGVS descriptions and the transcript. For example, in PMID 9535554^33^, 5 of 6 cases matched via the unchanged p.Arg58Gln descriptor. In contrast, blinded human reviewers flagged cases as problematic when HGVS expressions or transcripts were inconsistent. Together, these findings suggest that while hallucinated variant presence was uncommon in this constrained setting, future prompt design should require a fully concordant variant description.

### Model reasoning and justification

Model justifications helped audit each counting decision and were most informative where the phenotype had to be inferred. For *MYBPC3* c.1828G>C (p.Asp610His), PMID 32531501^34^, reported in a patient who also carried a pathogenic *GLA* variant, models had to map the free-text finding “Asymmetrical Obstructive DD grade II LGE in septum” to hypertrophic cardiomyopathy, the required phenotype, and attribute it to the correct gene. All twelve models did so and returned the correct count, several locating the variant in a footnote to a supplementary table of *GLA* variants. Justifications also exposed de-duplication reasoning: for CDH1 c.2100del a model recognized that two supplementary-table entries described the same family where the curator had counted two probands (Supplementary Example S6).

## Discussion

This study demonstrates that reasoning-capable LLMs can support automation of a key bottleneck in large-scale clinical variant curation: extracting literature-based case evidence supporting ACMG/AMP PS4 with ClinGen VCEP guidance. We observed more variation in model performance on tasks requiring that ClinGen guidance be applied, and detailed error analysis shows that most PS4 inaccuracies reflect guideline-execution failures (Supplementary Example S2).

A prompt baseline comparison showed the importance of prompt engineering, and specifically that models natively lack variant interpretation guidelines. Without careful prompting, gene-specific PS4 calculation is not reliably available to these models. This effect was most pronounced for the smallest model, Claude Haiku 4.5, which moved to within reach of the frontier group, indicating that prompt guidance can partly compensate for model scale.

These results move model selection from an accuracy question to an operational one. Five of six contemporary models could not be separated on PS4 case counting, including the open-weight GLM-5 model. Only the smallest model was statistically distinguishable. Among the tied models, cost per publication-variant pair and latency vary several-fold and are not correlated. Model selection therefore depends on cost, latency, governance and validation considerations.

Clinical deployment will remain constrained by residual errors including hallucinations, security, privacy and bias risks, and evolving regulatory expectations^35–40^. Ambiguity in existing case reports, especially phenotype descriptions and family structure, and increased run-to-run variability particularly with complex publications (Supplementary Text S2) means we will continue to require expert judgment. We therefore support a pragmatic hybrid model: LLMs apply guidelines to generate structured evidence summaries with citations and uncertainty flags, while experts adjudicate complex cases.

Literature-derived evidence remains a rate-limiting step in variant classification. It is likely that future revisions of the ACMG/AMP framework^41^ will increase this complexity. Today’s LLMs can absorb this load. Applied with the considerations set out above, reasoning-capable LLMs are becoming a practical requirement for doing variant classification at scale.

### Limitations

The truth set was drawn from historical HGSC-CL records, so these results describe this benchmark rather than the wider literature. The applicability of PS4 evidence itself varies by interpretation context; rare disease variants will likely have fewer publications than common adult-onset disease. Task 3 is a limited proof of concept and we would require a dedicated study to fully describe performance on that problem. Error categorization covered all six Phase 1 models but one lineage per vendor in Phase 2. Model reasoning depth was not uniform across providers.

## Data Availability

All data produced in the present work are contained in the manuscript and the GitHub repo at https://github.com/BCM-HGSC/LiteratureMining4VariantInterpretation

## Data sharing

The expert-curated truth set, including negative-controls, is provided as Supplementary Data files with this article. Aggregate model outputs, negative-control results, and analysis results are publicly available at https://github.com/BCM-HGSC/LiteratureMining4VariantInterpretation, along with the code used to generate all reported performance metrics, figures, and tables. Model prompts, output schema specifications, and configuration details are included in the Supplementary Appendix.

## Acknowledgements

We acknowledge Amazon Web Services (AWS) for providing the cloud infrastructure used in this work, including access to large language models through AWS Bedrock.

## Contributions

EV and BY conceived and designed the study. EV identified the study data and designed the benchmarking truth-set. BY, EV, SJ, SK, and CG created and manually validated the truth-set. MM, FD, and VM developed the study software and pipeline. MM, EV and AM performed the data acquisition, processing, statistical analysis, and benchmarking, and generated the performance metrics and figures. BY, EV, SJ, SX, and CG reviewed model results. MM and EV drafted the manuscript. RG provided overall supervision and administrative support for the study. All authors contributed to the interpretation of the results, critically reviewed and edited the manuscript, approved the final version to be published, and agree to be accountable for ensuring the integrity and accuracy of the work.

