## Supplementary Appendix for "Performance Characteristics of Reasoning Large Language Models for Evidence Extraction from Clinical Genomics Literature"

### Supplementary Table S1: Model information and inference parameters

|  | **Model** | **Release date** | **Model identifier** | **Invocation API** | **Temp.** | **Reasoning / thinking** | **Knowledge cutoff** |
| --- | --- | --- | --- | --- | --- | --- | --- |
| Phase 1 | Gemini 2.5 Pro^1^ | Jun 2025 | gemini-2.5-pro | Gemini API | 0 | thinking level = medium | Jan 2025 |
|  | GPT-5^2^ | Aug 2025 | gpt-5-2025-08-07 | OpenAI API | Not accepted | reasoning_effort = high | Sep 2024 |
|  | o3^3^ | Apr 2025 | o3-2025-04-16 | OpenAI API | Not accepted | reasoning_effort = high | Jun 2024 |
|  | o4-mini^3^ | Apr 2025 | o4-mini-2025-04-16 | OpenAI API | Not accepted | reasoning_effort = high | Jun 2024 |
|  | Claude Sonnet 4^4^ | May 2025 | anthropic.claude-sonnet-4-20250514-v1:0 | AWS Bedrock | 0 | extended thinking on |  |
|  | Claude Opus 4.1^4,5^ | Aug 2025 | us.anthropic.claude-opus-4-1-20250805-v1:0 | AWS Bedrock | 0 | extended thinking not enabled | Jan 2025 |
| Phase 2 | Claude Haiku 4.5^6^ | Oct 2025 | anthropic.claude-haiku-4-5 | AWS Bedrock | 0 | extended thinking not enabled | Feb 2025 |
|  | Gemini 3.1 Pro Preview^7^ | Feb 2026 | gemini-3.1-pro-preview | Gemini API | 0 | thinking level = high | Jan 2025 |
|  | Z.AI GLM-5^6^ | Feb 2026 | zai.glm-5 | AWS Bedrock | 0 | reasoning_effort = high |  |
|  | Claude Opus 4.8^8^ | May 2026 | anthropic.claude-opus-4-8 | AWS Bedrock | Not accepted | extended thinking not enabled | Jan 2026 |
|  | Claude Sonnet 5^9^ | Jun 2026 | anthropic.claude-sonnet-5 | AWS Bedrock | Not accepted | adaptive thinking = on; effort = high | Jan 2026 |
|  | GPT-5.6 Sol^10^ | Jul 2026 | openai.gpt-5.6-sol | AWS Bedrock | Not accepted | reasoning_effort = medium | Feb 2026 |

All phase 1 and phase 2 models were run with identical inputs, the same prompt template, and required to return a single JSON object conforming to the predefined output schema. Total input context was capped at 200,000 tokens. Model information and inference listed below:

### Supplementary Text S1: Prompt engineering and output schema

This section provides full details of the prompt engineering and schema design referenced in the main Methods. All models were evaluated using identical prompts and a fixed schema-constrained JSON output format to enable controlled comparisons and guideline-constrained PS4 case counting. The prompt instructed models to determine whether the target variant was reported and to identify and count PS4-eligible probands in each publication by applying the gene-specific PS4 digest based on ACMG-AMP and ClinGen VCEP guidelines. Models were instructed to base outputs only on the provided publication text and PS4 digest, to avoid unsupported inference, and to return schema-compliant outputs to minimize post-processing ambiguity. Prompt and schema development were iteratively refined before final benchmarking. The primary benchmark used the finalized v7 prompt and schema configuration. Prompt S0 is invoked with the LLM Gemini 2.5 Pro to generate the PS4 guidelines digest using ACMG-AMP and ClinGen VCEP guidelines. Prompt S1 utilizes Prompt S0 and is invoked with each of the evaluated models to generate results for tasks 1 and 2. See below for details:

#### Prompt S0: PS4 guideline-digest generation (ClinGen VCEP + ACMG/AMP)

**Purpose:** Before evidence extraction, we generated a PS4 “digest” that converts ACMG/AMP guidance and ClinGen VCEP gene-specific text into a PS4-only, citation-anchored rubric (YAML). The digest explicitly determines ps4_mode (*case_control*, *proband_count*, or *not_applicable*), enumerates inclusion/exclusion rules (including inheritance and independence/duplication handling), and captures any numeric thresholds verbatim for downstream rule-constrained extraction.

**Inputs:**

- **{gene}:** Gene symbol.
- **{gene_txt}:** Parsed ClinGen VCEP gene-specific specification text (verbatim). We generated PS4 guideline digests for all genes with ClinGen VCEP specifications that were released or designated as ready for release at the time of analysis.
- **{acmg_text}:** Parsed generic ACMG/AMP PS4 guidance text (verbatim).

**Prompt (verbatim template):**

You are a variant curation assistant credentialed by the American Board of Medical Genetics and Genomics.
Summarize PS4-relevant rules using only the provided ClinGen VCEP gene-specific text {gene_txt} and the generic ACMG/HGSC text {acmg_text}.
Do NOT invent or use external information. Cite line/table numbers for every numeric threshold.

** Determine PS4 mode from the text (gene-specific overrides ACMG; prefer proband counting when allowed): **
1) not_applicable — If the gene-specific text says PS4 is not used/applicable, set ps4_mode: "not_applicable" and summarize the alternative evidence path the VCEP specifies (e.g., tumor IHC → PP4; calibrated functional/LR paths), with citations from {gene_txt}.
2) case_control — Else, if the gene-specific text forbids proband/family counting for PS4 or provides a case–control OR/LR/RR framework, set ps4_mode: "case_control" and summarize required statistics (OR/RR/LR, CI, p-value) and any dataset-matching constraints, with citations from {gene_txt}.
3) proband_count — Else, if the gene-specific text provides a proband/family counting rubric (e.g., phenotype-point system; one case per family), set ps4_mode: "proband_count" and list the counting/point thresholds with citations from {gene_txt}.
4) Default (proband_count via ACMG) — If none of the above appear in {gene_txt}, default to proband_count using the generic ACMG PS4 description from {acmg_text} (e.g., multiple unrelated affected probands with absence in controls).

Focus on:
- PS4 criteria, thresholds, case/family counting rules, phenotype/inheritance constraints, and all numeric thresholds (cite each).
- When to use PM3 or other alternatives (e.g., recessive cases, compound hets) instead of PS4 (cite).
- All exceptions, gene-specific modifiers, and details for independence/de-duplication. If {gene_txt} is silent on independence, use {acmg_text} for the definition and cite it.
- Every numeric threshold or criterion must have an explicit citation (e.g., "Minimum 3 cases (Table 2)").

Your output must match this YAML schema:
gene: "{gene}"
guideline_version: |
 - ClinGen VCEP version ← cite line / table
 - ACMG version ← cite
ps4_mode: "proband_count | case_control | not_applicable"
ps4_mode_rationale: |
 <1–3 sentences explaining why this mode was chosen; MUST cite exact lines/tables from the provided text>
ps4_rules:
 inclusion_criteria:
 exclusion_criteria:
 case_thresholds:
 independence_definition:
 zygosity_notes:
 non_ps4_recommendations:
variant_classification:
 key_criteria:
 gene_specific_modifiers:
inheritance:
 mode: "Autosomal Dominant | Recessive | X-linked"
 penetrance_notes:
 special_cases:
phenotypes:
 allowed:
 - name: "<phenotype 1>"
 inheritance: "<Dom/Rec/...>"
 source: "<cite>"
citations: []

**Example S0 output (generic ACMG/AMP default; abbreviated):**

When no gene-specific specification is provided, S0 defaults to a generic ACMG/AMP PS4 digest, including the proband-count interpretation and the distinction between proband counting vs formal case-control PS4 statistics.

ps4_mode: "proband_count"

ps4_mode_rationale: >

Default interpretation of generic ACMG/AMP; “multiple unrelated patients” and “absent in controls”

ps4_rules:

inclusion_criteria:

- Variant observed in multiple unrelated patients with the same phenotype (Table 3, PS4 Note 2).

exclusion_criteria:

- Variant must be absent in controls (Table 3, PS4 Note 2).

case_thresholds:

- No numeric threshold for “multiple” is specified (Table 3, PS4 Note 2).

independence_definition:

- Cases must be unrelated; avoid duplicate counting across overlapping publications (Table 3, PS4 Note 2; Literature/Database Use).

**Example S0 output: PS4 proband_count mode (gene-specific ClinGen VCEP digest - APC; abbreviated)**

Illustrates a gene in which the VCEP permits **proband/family counting** for PS4 (rather than restricting PS4 to formal case-control statistics). The output below is the structured GUIDE (YAML) produced by Prompt S0 and passed verbatim into the downstream extraction prompt (Prompt S1).

gene: "APC"

guideline_version:

- ClinGen InSiGHT Hereditary Colorectal Cancer/Polyposis VCEP Specifications v2

- ACMG/AMP 2015 Standards and Guidelines

ps4_mode: "proband_count"

ps4_mode_rationale: >

APC uses a phenotype-based point system from unrelated probands (PS4 section; Table 1).

ps4_rules:

inclusion_criteria:

- Max 1 point per proband; points assigned by phenotype category (Table 1).

case_thresholds:

- PS4_Supporting: 1–1.5 points; PS4_Moderate: 2–3.5; PS4_Strong: 4–15.5; PS4_Very Strong: ≥16.

independence_definition: >

Count per proband; max 1 point per proband to prevent within-family double counting (Table 1).

#### Prompt S1: v7 prompt and schema for Task 1 - variant detection and Task 2 - PS4 case counting

**Purpose.** Prompt S1 operationalizes publication-level evidence extraction and PS4-eligible case counting using (i) the publication text, (ii) the gene-specific PS4 guideline digest generated by Prompt S0 and (iii) provided instructions. The prompt instructs models to (1) determine whether the target variant is reported in the publication (Task 1) and (2) count PS4-eligible probands under the PS4 mode and rules specified in the digest (Task 2). To enable controlled comparisons across models and reduce post-processing ambiguity, outputs were required to conform to a fixed schema-constrained JSON format (Schema S1), with explicit citations supporting all eligibility decisions.

**Inputs:**

- **{guide}:** Gene-specific PS4 digest (Prompt S0 output).
- **{document_text}:** Parsed publication text to be searched (including supplementary text when available).
- **{pmid}, {title}, {authors}, {year}:** Publication metadata.
- **{gene}, {tr}:** Gene symbol and transcript, if available.
- **{vlist}:** Target variant list (as provided to the pipeline).
- **{phenos}:** Phenotype dictionary used to define the closed set of allowed phenotypes.

**Prompt and output schema S1:**

You are an ABMGG-credentialed clinical laboratory geneticist and an expert in AMP/ACMG + ClinGen guidance. Your task is to perform a rigorous, auditable extraction of data for the PS4 criterion. Use ONLY the provided GUIDE and DOCUMENT. {guide}

PMID="{pmid}"; TITLE="{title or ''}"; AUTHORS={authors or []}; YEAR="{year or ''}"; GENE="{gene}"; TRANSCRIPT="{tr}"; VARIANT_LIST={vlist}; PHENOTYPE_DICT={phenos}; ALLOWED_PHENOTYPES = A definitive, closed list constructed ONLY from the PHENOTYPE_DICT and any phenotypes explicitly named as included in the provided GUIDE.

DOCUMENT (the publication to search) {document_text}

**CARDINAL RULE: COUNT FIRST, EVALUATE LATER** Your primary mission is to **count individuals** based on their reported phenotype and variant status in BOTH case series and case control studies. You are **NOT applying the final PS4 strength criterion**.

STUDY-TYPE GATE (apply before any counting)

- Classify the DOCUMENT as one of: • primary: authors present genotyping/sequencing or case ascertainment in case series or case control publications. • secondary-only: methods/assay validation, literature review, previously reported study, meta-analysis, functional study, registry/database re-tabulation (e.g., ClinVar, gnomAD, UK Biobank) with no new individual-level clinial cases. Rules:
- If secondary-only → set: a=b=c=d=0, total_ps4_case_counts=0 Then STOP. Record logic in counting_justification and 'model_reasoning`.

*GOALS (counting-only)*

1. Set ps4_mode from GUIDE (case_control | proband_count | not_applicable) and cite the exact GUIDE place.
2. Find the variant (or declare not found). If variant not found → a=b=c=d=0; total_ps4_case_counts=0.
3. Only tally counts per mode (do NOT apply thresholds or compute stats). Copy any GUIDE thresholds verbatim into ps4_thresholds_applied for reference only.
4. If ps4_mode=case_control → build a 2×2 (a,b,c,d). If the paper reports only aggregates, **estimate counts** per Estimation Policy below. Do NOT assign PS4 strength; enumerate eligibility only.
5. If ps4_mode=proband_count → identify PS4-eligible unrelated probands from affected+variant-positive (bucket “a”). Do NOT assign PS4 strength; enumerate eligibility only.
6. If ps4_mode=not_applicable → do not count (a,b,c,d) or PS4; record the alternate path from GUIDE.
7. Record all exclusions, duplicates/overlaps, aggregates/derived counts, inheritance edge cases, and citations.
8. Output EXACT JSON per the schema (no extra keys, no prose).

*Perform all evaluations strictly from the provided input, publication 'DOCUMENT' and the supplied 'GUIDE', following tasks listed below. Do not hallucinate or make assumptions beyond the provided data. *Provide citations for any guidelines or standards you reference in your reasoning.

GLOBAL DO/DON’T

- DO dedupe relateds/overlaps; one proband per family unless independence is explicit.
- DON’T compute OR/CI/p.
- DON’T apply or infer PS4 strength; DON’T zero counts because a threshold (e.g., “multiple probands”, OR≥X) isn’t met.
- Affected = ALLOWED_PHENOTYPES only(quote & cite). Controls require explicit healthy/unascertained language.
- 2x2 definitions: a = affected + variant positive b = affected + variant negative c = unaffected + variant positive d = unaffected + variant negative
- “Affected” includes unrelated cases matching ALLOWED_PHENOTYPES (explicit or clearly inferred and quoted).
- Default assumption of relatedness: Unless the authors explicitly indicate that cases are unrelated, assume they are related and count them as one case.
- “Unaffected” requires an explicit healthy/control statement; if unclear, do not treat as unaffected.
- Never mix proband counts with case-control metrics in the same total.

ESTIMATION POLICY (for counts only; never for OR/RR/HR)

- If the paper gives frequencies/percentages and denominators, derive integer counts (nearest whole; 0.5 up) and document quotes and arithmetic.
- Affected-only catalog/table minimum: if variant is listed among affected cases with no per-variant count, set a=1 (conservative minimum) unless a defensible higher estimate is provided; record both.
- Controls optional: populate b/c/d only when reported or derivable; otherwise set to 0 with explanation.

**Structured Reasoning Workflow** (INTERNAL STEP 1: Foundational Analysis) … (as specified) (INTERNAL STEP 2: Self-Critique & Refinement) … (as specified) (INTERNAL STEP 3: Final Output Generation) … (as specified)

**OUTPUT: ( JSON only, no comments, no extra keys)**

```json

{{

"schema_version": "ps4_v7_reported",

"ps4_mode": "<case_control | proband_count | not_applicable>",

"ps4_mode_citation": "<exact GUIDE section/table>",

"ps4_thresholds_applied": "<verbatim thresholds/rubric from GUIDE, or 'generic ACMG PS4 in GUIDE'>",

"publication": {{

"pubmed_id": "{pmid}",

"title": "{title or ''}",

"authors": {authors or []},

"year": "{year or ''}"

}},

"variant_metadata": {{

"gene": "{gene}",

"variant_list": "<comma-separated as provided>",

"transcript": "{tr}",

"phenotypes_considered": "<ALLOWED_PHENOTYPES>"

}},

"variant_found": true | false,

"variant_forms": ["<exact strings seen in DOCUMENT>"],

"a": <int>, "b": <int>, "c": <int>, "d": <int>,

"total_ps4_case_counts": <int>,

"mode_sections": {{

"case_control": {{"case_control_data": "<case_control_data if available, else empty string>"}},

"proband_count": {{

"unrelated_probands": <int>,

"probands": [{{"id":"FamX/PrY","phenotype":"<text>","independent":true,"zygosity":"<het|hom|compound_het|NA>","points": null,"source":"<page/table/figure>"}}],

"rubric_source": "<GUIDE table/section or 'NA'>"

}},

"not_applicable": {{"alternate_path": "<verbatim note from GUIDE>"}}

}},

"non_ps4_summary": [{{"label":"phenotype_unknown","count":0,"note":"", "source":""}}],

"counting_justification": "<The justification for all counts, including inclusion/exclusion criteria, deduplication logic, and estimation methods. This should be the final, corrected version.>",

"flag_for_review": false,

"citations": {{"guide": ["<…>"], "document": ["<…>"]}},

"model_reasoning": "<Illustrate the entire reasoning process, including the outcome of the self-critique step, explaining how the final determinations were made.>"

}}

```

#### Prompt S2: v0 prompt and schema for Task 1 - variant detection and Task 2 - PS4 case counting

**Purpose.** To estimate the contribution of the domain-specific instructions supplied in v7, we constructed a minimal task-only direct-call baseline, designated **v0**. For each publication–variant pair, v0 received the same publication text and basic identifying inputs as v7: gene, transcript, variant representation, phenotype, and PMID. It used the same pipeline, model, inference configuration, and structured-output parser as v7. Thus, the comparison was a paired prompt-ablation experiment.

The v0 prompt omitted the gene-specific ClinGen guideline digest and the principal domain-guidance components of v7, including the study-type gate, phenotype-list construction, case-estimation policy, family-level deduplication rules, inheritance-specific instructions, and self-critique step. It asked only whether the variant appeared in the publication and how many cases counted toward PS4 per ACMG/AMP guidance. A minimal JSON output contract was retained so that v0 and v7 outputs could be parsed and scored using the same deterministic analysis pipeline.

**Inputs:**

- **{document_text}:** Parsed publication text to be searched (including supplementary text when available).
- **{pmid}, {title}, {authors}, {year}:** Publication metadata.
- **{gene}, {tr}:** Gene symbol and transcript, if available.
- **{vlist}:** Target variant list (as provided to the pipeline).
- **{phenos}:** Phenotype dictionary used to define the closed set of allowed phenotypes.

**Prompt and output schema S2:**

You are a clinical geneticist. Read the publication below and answer two questions

about one variant.

GENE: {gene}

TRANSCRIPT: {tr}

VARIANT: {vlist}

PHENOTYPE: {phen}

PMID: {pmid}

1. Does this variant appear in this publication?

2. How many cases in this publication count toward the ACMG/AMP evidence code PS4 for this

variant and this phenotype?

<PUBLICATION>

{document_text}

</PUBLICATION>

Answer with JSON only, no prose outside it:

{{

"variant_found": true | false,

"total_ps4_case_counts": <integer>,

"counting_justification": "<one or two sentences: what you counted and why>",

"model_reasoning": "<one or two sentences: how you identified the variant and counted cases>",

"citations_guide": "<cite where and how you found the evidence>"

}}

### Supplementary Text S2: **Reproducibility (stochasticity) analysis**

**Methods**: To assess run-to-run variability (stochasticity)^11^ in model outputs, we executed each Phase 1 model - Gemini 2.5 Pro, GPT-5, o3, o4-mini, and Claude Sonnet 4 - using the primary v7 pipeline configuration - 10 times on a subset of 20 publications. Publications were manually stratified based on publication characteristics and curation difficulty identified during truth-set construction: *low-complexity* (n=10) publications typically described single cases with direct variant mentions and unambiguous case and phenotype descriptions, whereas *high-complexity* (n=10) publications typically included multiple cases, were longer and/or accompanied by supplements, used heterogeneous nomenclature, and more often required adjudication of phenotype relevance, case counting, or relatedness. Reproducibility was quantified as the proportion of runs matching the modal output for (1) Task 1 variant-detection calls and (2) Task 2 PS4 case counts. We assessed semantic consistency of the model’s reasoning outputs by computing pairwise cosine similarity of OpenAI text-embedding-3-large embeddings of the justification text across runs.

**Results**: Reproducibility varied across the heuristically defined low- and high-complexity publication subsets (Figure S1). In the low-complexity subset (Figure S1.A), reproducibility was high for variant detection (Task 1, 0.91-1.00) and PS4 case counting (Task 2, 0.96-1.00), with high reasoning-justification similarity (0.84-0.94). In the high-complexity subset (Figure S1.B), reproducibility decreased across all models, with wider variation in Task 1 (0.70-1.00) and Task 2 (0.77-0.94) and lower reasoning-justification similarity (0.79-0.91). The lower-capability models (o4-mini and Claude Sonnet 4) exhibited the greatest instability; the lowest Task 1 stability was observed for Claude Sonnet 4 (0.70), whereas the lowest Task 2 stability was observed for o4-mini (0.77).

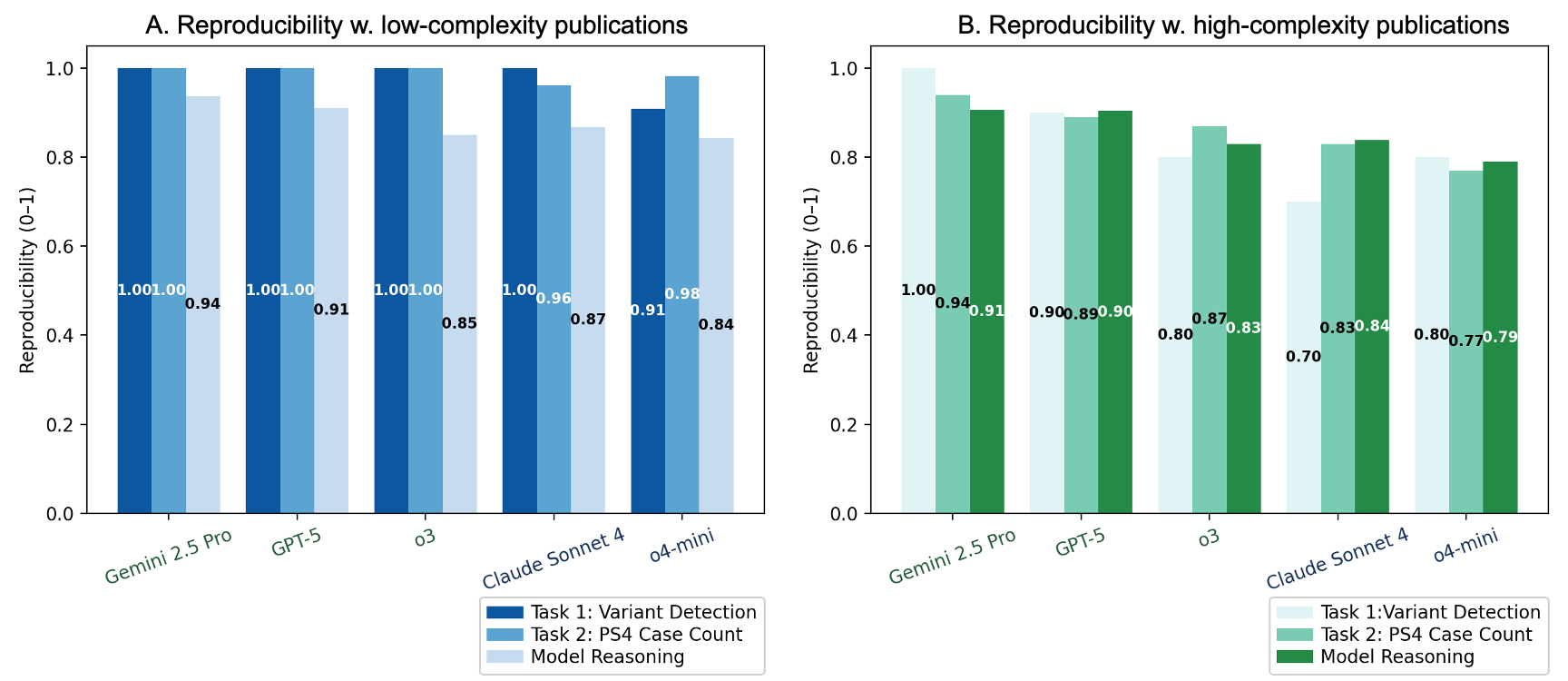

**Figure S1: Run-to-Run Reproducibility (Stochasticity) across low- and high-complexity publications.** Each Phase 1 model was run 10 times on 20 publications heuristically stratified as low complexity (n = 10) or high complexity (n = 10) based on manual review during truth-set construction; low-complexity publications (S1.A) typically described single cases with direct variant mentions and unambiguous case and phenotype descriptions, whereas high-complexity publications (S1.B) typically included multiple cases, had longer narratives and/or supplements, and used heterogeneous variant nomenclature, more often requiring adjudication of variant normalization, phenotype relevance, case counting, or relatedness. For Task 1 (variant detection) and Task 2 (PS4 case counting), reproducibility is reported as the proportion of runs matching the modal output for each publication (higher values indicate less stochastic variation). For reasoning justifications, semantic consistency is summarized as the mean pairwise cosine similarity of text embeddings across runs (0-1). Reproducibility was generally higher in the low-complexity subset and declined in the high-complexity subset, with greater variability among the lower-capability models (o4-mini and Claude Sonnet 4).

### Supplementary Text S3: Model error categories

**Purpose**: To profile failure modes in Task 2 (PS4 case-count extraction), we manually reviewed all non-concordant publication-variant pairs (predicted count ≠ truth-set count). Each discordant output was assigned one primary error category from a prespecified taxonomy (below). Categories were defined to reflect the most proximal cause of miscounting (e.g., incorrect relatedness/independence handling vs incorrect guideline logic. When multiple issues were present, the primary category was assigned based on the earliest decision point that would have corrected the final count (e.g., variant not found → phenotype mapping → relatedness/independence and deduplication → guideline logic). If no single category could be confidently assigned, errors were labeled Unknown.

**Error categories (primary labels)**

- **Relatedness - Overcount:** The model counted family members when it should have counted only the proband: nonconservative handling of relatedness or independence, treating related individuals from the same family as independent cases.
- **Relatedness - Undercount:** The model treated cases as possibly from the same family, often because the publication gives too little information to rule relatedness out, even though they may in fact be unrelated. Overly conservative handling of relatedness or independence.
- **Variant not found:** Failed to identify or correctly map the prespecified variant (including missed aliases/representations), resulting in incorrect counting.
- **Phenotype association / mismatch:** Included or excluded cases due to mismatch with the allowed PS4 phenotype set, or unresolved phenotype ambiguity in the publication.
- **Inheritance / zygosity error:** Misclassified inheritance context or zygosity (e.g., heterozygous vs homozygous vs compound heterozygous; sex-linked context), including failures to route recessive/compound-heterozygous evidence to PM3-preferred handling when specified.
- **Guideline interpretation error:** Misapplied PS4 mode or eligibility rules from the ClinGen VCEP/ACMG-AMP digest and/or prompt instructions (e.g. missed eligible cases, including failure to extract or derive cases present in the publication; counting in not_applicable contexts such as for MSH6 gene; assigning PS4 strength when only a tally was requested; or misclassifying PM3-preferred contexts as PS4-eligible).
- **Duplicate case:** The model counted the same person more than once, because the publication indicated prior reporting or an overlapping cohort.
- **Not a primary case report:** Treated secondary-only sources (e.g., review/meta-analysis, database tabulation, functional/assay validation without new individual-level clinical cases) as case evidence.
- **Primary case not recognized:** Failed to recognize bona fide case evidence in a primary publication and therefore did not count it.
- **Missed evidence:** Relevant counts were present in the provided text/tables/supplements but were not used in the model’s tally.
- **Unknown:** discordance could not be confidently attributed to one category.

### Supplementary Table S3: Task 1 (Variant Detection): Confusion Matrix Counts and Performance Metrics for LLMs Versus Expert-Curated Truth-set

Purpose: For all phase 1 and phase 2 models, across 281 publication-variant pairs (275 unique publications), we evaluated each model’s ability to determine whether a prespecified variant was reported in the publication, using an expert-curated truth-set reference. Results are summarized as confusion-matrix counts and derived performance metrics.

| **Phase** | **Model** | **N** | **TP** | **FP** | **FN** | **TN** | **Accuracy** | **Sensitivity** | **Specificity** | **PPV** | **NPV** | **F1** |
| --- | --- | --- | --- | --- | --- | --- | --- | --- | --- | --- | --- | --- |
| Phase 1 | GPT-5 | 281 | 264 | 0 | 5 | 12 | 0.982 | 0.981 | 1.000 | 1.000 | 0.706 | 0.991 |
| Phase 1 | Gemini 2.5 Pro | 281 | 262 | 0 | 7 | 12 | 0.975 | 0.974 | 1.000 | 1.000 | 0.632 | 0.987 |
| Phase 1 | o3 | 281 | 262 | 0 | 7 | 12 | 0.975 | 0.974 | 1.000 | 1.000 | 0.632 | 0.987 |
| Phase 1 | Opus 4.1 | 281 | 258 | 0 | 11 | 12 | 0.961 | 0.959 | 1.000 | 1.000 | 0.522 | 0.979 |
| Phase 1 | Sonnet 4 | 281 | 257 | 3 | 12 | 9 | 0.947 | 0.955 | 0.750 | 0.988 | 0.429 | 0.972 |
| Phase 1 | o4-mini | 281 | 250 | 0 | 19 | 12 | 0.932 | 0.929 | 1.000 | 1.000 | 0.387 | 0.963 |
| Phase 2 | Opus 4.8 | 281 | 268 | 0 | 1 | 12 | 0.996 | 0.996 | 1.000 | 1.000 | 0.923 | 0.998 |
| Phase 2 | Sonnet 5 | 281 | 267 | 1 | 2 | 11 | 0.989 | 0.993 | 0.917 | 0.996 | 0.846 | 0.994 |
| Phase 2 | GPT-5.6 | 281 | 267 | 2 | 2 | 10 | 0.986 | 0.993 | 0.833 | 0.993 | 0.833 | 0.993 |
| Phase 2 | Gemini 3.1 Pro Preview | 281 | 266 | 1 | 3 | 11 | 0.986 | 0.989 | 0.917 | 0.996 | 0.786 | 0.993 |
| Phase 2 | GLM-5 | 281 | 263 | 1 | 6 | 11 | 0.975 | 0.978 | 0.917 | 0.996 | 0.647 | 0.987 |
| Phase 2 | Haiku 4.5 | 281 | 258 | 0 | 11 | 12 | 0.961 | 0.959 | 1.000 | 1.000 | 0.522 | 0.979 |

**Legend (counts):**

**N**: total publication–variant pairs evaluated

**TP (true positive):** model indicates the variant is reported **and** ground truth confirms presence

**TN (true negative):** model indicates the variant is not reported **and** ground truth confirms absence

**FP (false positive):** model indicates the variant is reported but ground truth indicates **absence** (hallucinated presence)

**FN (false negative):** model indicates the variant is not reported but ground truth indicates **presence** (missed presence)

**Metric definitions:**

**Accuracy:** proportion of all predictions that were correct = (TP+TN)/N

**Sensitivity (recall, true-positive rate):** proportion of true variant-positive publications correctly identified = TP/(TP+FN)

**Specificity (true-negative rate):** proportion of true variant-negative publications correctly

excluded = TN/(TN+FP)

**PPV (precision, positive predictive value):** proportion of model-positive calls that were truly positive = TP/(TP+FP)

**NPV (negative predictive value):** proportion of model-negative calls that were truly negative = TN/(TN+FN)

**F1 score:** harmonic mean of precision and recall = 2⋅(PPV⋅Sensitivity)/(PPV+Sensitivity)

### Supplementary Example S1: Legacy nomenclature can affect Task 1 variant detection for smaller, efficiency oriented models

**Purpose:** Illustrates a Task 1 false negative in o4-mini and Claude Sonnet 4, driven by non-standard/legacy variant representations: the publication reports the prespecified variant using legacy mature-protein numbering and a historical cDNA coordinate, which o4-mini and Claude Sonnet 4 fail to map to the input representation.

**Input**: PMID 10559517^12^**;** LDLR**;** NM_000527.5(LDLR)**;** p.Asp482His**;** c.1444G>C
Legacy representation in publication: “D461H” and “1443 G/C”

Location of variant evidence: Table 1

**Model outputs (excerpt from counting_justification):**

- **o4-mini (variant not found):**
  “An exhaustive search of the DOCUMENT, including Table 1 listing all LDLR mutations, found no mention of p.Asp482His or c.1444G>C. Therefore variant_found=false and no cases were tallied.”
- **Claude Sonnet 4 (variant not found):**
  “Created exhaustive alias list for p.Asp482His (c.1444G>C) including: D482H, Asp482His, D461H (legacy numbering accounting for signal peptide), 1444G>C, 1443G>C (legacy), IVS notation if applicable. Searched entire document for all aliases. The variant of interest was not found in any form throughout the publication.”
- **GPT-5 (variant found; legacy mapping):**
  “Variant identification and alias reconciliation: Input variant is LDLR p.Asp482His (c.1444G>C) on NM_000527.5. In this paper, the same molecular change is reported using legacy mature-protein numbering and a slightly different cDNA coordinate: D461H with base change 1443 G/C (Table 1, page 312; mis-sense discussion page 315). LDLR mature-protein numbering is offset by 21 amino acids relative to full-length numbering; thus p.D461H (mature) corresponds to p.D482H (full-length). The cDNA coordinate discrepancy (1443 vs 1444) is consistent with transcript/version-dependent c. numbering used historically; functionally, D→H arises from a first-base G→C change in the Asp codon, matching 1443 G/C in this paper and widely known mappings for D482H. Therefore, we treat D461H (1443 G/C) in this document as the same event as p.Asp482His (c.1444G>C) in the input. Exact strings observed: 'D461H' and '1443 G/C' (Table 1).”
- **Gemini 2.5 Pro (variant found; legacy mapping):**
  “The variant of interest, p.Asp482His (c.1444G>C), is identified in the publication using a legacy protein nomenclature, D461H, which accounts for the cleavage of the 21-amino acid signal peptide (482 - 21 = 461). The paper reports this variant as '1443 G/C', which is treated as the same molecular event.”
- **o3 (variant found; legacy mapping):**
  “Variant alias reconciliation: The input variant p.Asp482His (c.1444G>C, NM_000527.5) is historically numbered as D461H when the 21-residue signal peptide is excluded; Table 1 lists “D461H 1443 G/C”. This is the same molecular event and was therefore counted.”

**Why this is a nomenclature error:** The variant is present in the publication under legacy notation (“D461H; 1443 G/C”). In this example, o4-mini and Claude Sonnet 4 did not reconcile the legacy representation to the prespecified input (“p.Asp482His; c.1444G>C”) and therefore labeled the variant as absent, whereas GPT-5, Gemini 2.5 Pro and o3, mapped the legacy representation and correctly detected the variant.

### Supplementary Example S2: Guideline-execution failure for Task 2 PS4 proband case count

**Purpose:** Illustrates a Task 2 guideline-execution error in which models incorrectly apply the SLC6A8 PS4 rubric per VCEP guidelines and return a zero PS4 count**,** missing a truth-set PS4-eligible male case in the publication.

**Input:** PMID 33164824^13^; SLC6A8; p.Pro554Leu; c.1661C>T

Location of case evidence: Section 3.2 and Supplementary Table 1

**Model outputs (excerpts from counting_justification):**

- **Gemini 2.5 Pro (returned PS4 count = 0):** The publication reports a family with two affected siblings. The variant of interest, SLC6A8 c.1661C>T (p.Pro554Leu), was identified in the affected sister (Patient II:2) and her mother (Patient I:1). Patient II:2 has a confirmed diagnosis of 'cerebral creatine deficiency syndrome 1', which matches the allowed phenotype 'creatine transporter deficiency'. This individual is counted in 'a'. The affected brother (Patient II:1) does not have the variant and is counted in 'b'. The mother (Patient I:1) has the variant, but her diagnosis of CCDS1 is not confirmed, and required biochemical data is missing, so she is not counted. The provided GUIDE specifies `ps4_mode` as `proband_count` with strict inclusion criteria for the SLC6A8 gene: 'Probands must be independent males' and 'must have an elevated urine creatine/creatinine ratio on at least one occasion'. Patient II:2, the only individual with the variant and confirmed disease in this report, is female and does not have an elevated urine creatine/creatinine ratio (as per Supplementary Table 1). Therefore, this individual does not meet the VCEP's specific criteria for inclusion in the PS4 count. The final PS4 proband count from this paper is 0.
- **GPT-5 (returned PS4 count = 0):** Study-type gate: Primary report with new exome sequencing and clinical/biochemical data on a family; eligible for counting. Variant identification: The paper explicitly reports SLC6A8 (NM_005629.4) c.1661C > T, p.(Pro554Leu) with genomic coordinate “hg19, chrX:g.152960238C > T,” present in the heterozygous mother (I:1) and daughter (II:2), and absent in the male sibling (II:1) and father (I:2) (Results 3.2). We captured all exact strings used in the document. Phenotype mapping: The daughter II:2 is diagnosed with Cerebral Creatine Deficiency Syndrome 1 (CCDS1), synonymous with creatine transporter deficiency (Introduction/Discussion; Results 3.2), which is within the allowed phenotype scope. PS4 application mode: Per the SLC6A8 VCEP, PS4 is a proband-counting system restricted to independent MALE probands with an elevated urine creatine/creatinine ratio at least once, and the variant must meet PM2_Supporting. In this report, there are no variant-positive males. The only variant-positive affected individual is a heterozygous female (II:2). Her urine creatine values (mmol/mol creatinine) at 28 months (846; ref 28–1700) and 8 years (948; ref 28–1700) were within reference ranges (Laboratory testing table), and regardless, females are not counted for PS4 under the VCEP. Therefore, a=0. No case-control cohorts are presented; b/c/d remain 0. Deduplication: A single family is reported; no unrelated probands. Literature-referenced prior male cases are not counted here because they are not primary data in this document. Note: Per PP4 additional specifications, an individual used for PP4 cannot also be included in the PS4 count; the female proband would be reserved for PP4 if used and is excluded from PS4 in any case.

**Why this is a guideline-execution error:** Under the truth-set adjudication, this publication contains a PS4-eligible male proband meeting the SLC6A8 PS4_Supporting rubric, so the PS4 case tally should be non-zero (≥1)**.** The models’ failure reflects incorrect rubric execution (eligibility determination under the gene-specific PS4 constraints) rather than failure to locate the variant or phenotype.

### Supplementary Example S3: Claude Sonnet 4 relatedness-driven PS4 overcount

**Purpose**: Illustrates a Task 2 relatedness/independence overcount: the publication reports the variant at the family level (6 families; 12 patients), but Claude Sonnet 4 uses 12 individuals as the PS4-eligible case count under proband counting, inflating the tally relative to the truth-set rule (one independent proband per family unless individual-level independence is explicit).

**Input**: PMID 17024664^14^**;** VHL**;** p.Asn78Ser**;** c.233A>G *(legacy: c.446A>G)*
Location of case evidence: Supplementary Table S1

**Model output (Claude Sonnet 4; excerpt from counting_justification):**

“The variant p.Asn78Ser (c.233A>G in current nomenclature, c.446A>G in original nomenclature) was found in Supplementary Table S1. The table shows this variant occurred in 6 families with 12 total patients. Since the table presents data by families, these represent 6 unrelated probands with 12 total affected individuals. For PS4 proband counting, we count the number of affected individuals (12) rather than just probands, as each affected individual with the variant contributes to the evidence. All cases have Von Hippel-Lindau syndrome, which matches the allowed phenotypes. The study is autosomal dominant inheritance as stated in the guide.”

**Why this is an overcount:** Because the evidence is family-aggregated, counting affected individuals as independent PS4 cases overstates evidence under the truth-set’s one-proband-per-family deduplication rule.

### Supplementary Example S4: Gemini 2.5 Pro Duplicate Case

**Purpose**: Illustrates the instance of a duplicate case, in which the same individual from a primary publication was incorrectly counted as a case in this publication, distinguished from the more common error of counting related individuals as independent probands.

**Input**: PMID 31463198^15^; MT-TF; m.586G>A

Location of case evidence: Supplementary Table S1

**Model output (Gemini 2.5 Pro; excerpt from counting_justification):**

“The publication is a primary case report. The variant m.586G>A was identified in two unrelated individuals with phenotypes consistent with 'mitochondrial disease'. Case 1 is a 14-year-old boy, newly reported in this document, with 'non-progressive muscle weakness and myalgia' and 'mitochondrial myopathy' (Page 1, Para 2, 4). Case 2 is a 57-year-old woman previously reported by Young et al., 2010 (Ref [5]), with a 'progressive neurodegenerative disorder' (Page 1, Para 6). The authors of this paper explicitly establish the two cases as unrelated by presenting them as distinct observations from different sources and publications. Therefore, two independent probands are counted for PS4. No control cohort was described, so b, c, and d are 0.”

**Why this is a duplicate:** Here a single individual from another primary publication was counted twice, so the error is Duplicate case

### Supplementary Example S5: Effect of prompt ablation

**Purpose**: To estimate the contribution of guideline informed v7 prompt on Task 1 and Task 2 performance, by comparing with the v0 prompt, a minimal task-only baseline (Supplementary Text S2).

The same 281 publication–variant pairs, models and inference configuration were re-run under v0, which supplies only the gene, transcript, variant, phenotype and publication text, omitting the gene-specific ClinGen guideline digest, the study-type gate, phenotype-eligibility rules, and the inheritance and relatedness screens present in v7. Full v0 and v7 prompt templates and output schemas are given in Supplementary Text S1 and S2 and are not repeated here.

Three genes in the benchmark - MLH1, MSH6, and PMS2 - have ClinGen Variant Curation Expert Panel (VCEP) specifications that do not evaluate PS4 by proband counting. The benchmark contained 20 publication–variant pairs for these genes (MLH1, 7; MSH6, 11; PMS2, 2), with a curated PS4 case count of zero for every pair. These pairs provide a direct illustration of what occurred when the same publication and variant were evaluated using the v0 prompt versus the guideline-informed v7 prompt for the Phase 2 models (Supplementary Table S4).

Variant detection (Task 1) was not the limiting step: all six Phase 2 models identified the queried variant in all 20 pairs under both prompts (120/120 in each). Under v0, no model invoked the VCEP restriction; instead, the models returned a nonzero PS4 count on 13-17 of the 20 pairs, 278 cases in total against a curated zero. The 26 v0 evaluations that did return zero did so for publication-local reasons such as phenotype association issues, not because the model recognized that PS4 does not apply to the gene. Under v7, all six models correctly selected `ps4_mode = not_applicable` and returned zero for every pair.

Supplementary Table S4. Results in the PS4-not-applicable stratum

| **Phase 2 Models** | **v0 pairs with count >0** | **Sum of v0 PS4 counts** | **v7 pairs with count >0** | **Sum of v7 PS4 counts** |
| --- | --- | --- | --- | --- |
| Claude Opus 4.8 | 13/20 | 50 | 0/20 | 0 |
| Claude Sonnet 5 | 14/20 | 48 | 0/20 | 0 |
| Claude Haiku 4.5 | 17/20 | 36 | 0/20 | 0 |
| GPT-5.6 Sol | 17/20 | 54 | 0/20 | 0 |
| GLM-5 | 16/20 | 34 | 0/20 | 0 |
| Gemini 3.1 Pro | 17/20 | 56 | 0/20 | 0 |
| **Total** | **94/120** | **278** | **0/120** | **0** |

One pair illustrates why. MSH6 c.10C>T (p.Gln4Ter), reported in 11 French Canadian families (PMID 25318681^16^), is otherwise straightforward: the variant and its carriers are readily identifiable. Given only the task-only v0 prompt, every model instead counted the affected carriers described in the text as PS4-eligible — Claude Opus 4.8, for example, returned 15 — because it had not been told the MSH6 rule. Given the same publication and variant plus the v7 guideline digest, the same models set ps4_mode = not_applicable, cited the MSH6 VCEP specification (PS4 designated "Not Applicable"; tumor IHC data used under PP4 instead of proband counting), and returned zero — while their unscored audit fields still recorded the same 11 families as PP4-relevant evidence and one additional case as PM3-relevant. The evidence was preserved; only the PS4 count changed.

### Supplementary Text S4: Evaluation of Z.ai GLM-5’s reasoning level parameter

To evaluate the impact of the reasoning-effort parameter, we compared GLM-5 at high and medium effort on the same 281 publication-variant pairs, holding prompt, context and output schema constant. The gain was concentrated in Task 2, where high effort recovered 250 of 281 counts against 233 at medium, a difference of 6.05 percentage points (McNemar P = 0.021). Task 1 accuracy rose from 96.09% to 97.86% and did not differ significantly (P = 0.227). Higher reasoning effort therefore improved PS4 case counting for this model, at 5.4 times the latency (15.7 to 85.3 seconds per pair) and 1.4 times the cost. Future work is needed to establish whether this generalizes to other models and which error categories higher effort recovers.

### Supplementary Example S6: Truth-set correction

The model's capabilities are highlighted by several notable examples and helped resolve. For the *CDH1* variant NM_004360.5(*CDH1*):c.2100del (p.Val701SerfsTer21), referenced in PMID 26182300^17^, both human reviewers and the models successfully identified the variant in the publication. Initially, however, the manual review indicated two reported cases, while Gemini 2.5 Pro assessed it as a single case. During reconciliation, it was observed that the human reviewer assessed two cases because the variant appeared twice in the supplementary material with separate family IDs. The model, conversely, correctly determined that these represented the same family, likely by leveraging an author footnote and the lack of correspondence of variants reported in families with identical identifiers in the two table entries. This demonstrates the model's ability to detect subtle issues that might be overlooked by a human reviewer.

### References

1.	Gemini-2-5-Pro Model Card [Internet]. Available from: https://storage.googleapis.com/deepmind-media/Model-Cards/Gemini-2-5-Pro-Model-Card.pdf

2.	GPT-5 System Card [Internet]. Available from: https://arxiv.org/pdf/2601.03267

3.	OpenAI o3 and o4-mini System Card [Internet]. Available from: https://cdn.openai.com/pdf/2221c875-02dc-4789-800b-e7758f3722c1/o3-and-o4-mini-system-card.pdf

4.	System Card: Claude Opus 4 & Claude Sonnet 4 [Internet]. Available from: https://www-cdn.anthropic.com/6d8a8055020700718b0c49369f60816ba2a7c285.pdf

5.	System Card Addendum: Claude Opus 4.1 [Internet]. Available from: https://www-cdn.anthropic.com/9fa30625273bafdf5af82c93719d7ca606485a16/Claude%204.1%20System%20Card.pdf

6.	GLM-5-FP8 Model Card [Internet]. Available from: https://huggingface.co/zai-org/GLM-5-FP8

7.	Gemini 3.1 Pro Model Card [Internet]. [cited 2026 July 31];Available from: https://storage.googleapis.com/deepmind-media/Model-Cards/Gemini-3-1-Pro-Model-Card.pdf

8.	System Card: Claude Opus 4.8 [Internet]. Available from: https://www-cdn.anthropic.com/0f0c97ad20d8005706296bd92aa1c27c6b2f4f61/Claude%20Opus%204.8%20System%20Card.pdf

9.	System Card: Claude Sonnet 5 [Internet]. Available from: https://www-cdn.anthropic.com/283ef97c476cf442c91d9a37d5b214242a55bb92/Claude%20Sonnet%205%20System%20Card.pdf

10.	GPT-5.6 System Card [Internet]. Available from: https://deploymentsafety.openai.com/gpt-5-6/gpt-5-6.pdf
